# Change in incidence of cardiovascular diseases during the covid-19 pandemic and vaccination campaign: data from the nationwide French hospital discharge database

**DOI:** 10.1101/2022.08.01.22278235

**Authors:** Thierry Boudemaghe, Lucas Léger, Antonia Perez-Martin, Carey M. Suehs, Jean-Christophe Gris

## Abstract

**Importance:** The covid-19 pandemic induced a severe disruption in hospital activity. Cardiovascular illnesses represent a major health burden in industrialised countries and are second in terms of hospital bed occupancy in France. Considering the resources mobilized and the public health issue involved, it is necessary to study the impact of the pandemic on their incidences.

**Objective:** To monitor changes in the incidence of cardiovascular diseases during years 2020 and 2021 compared to 2019.

**Design:** Nationwide population-based cohort study.

**Setting:** French hospital discharge database between January 1 and October 30 in 2019, 2020 and 2021.

**Participants:** New patients hospitalized for vascular disease in Metropolitan France. A patient was considered as incident for a morbidity if not present in the database in the previous two years with the morbidity as the primary reason for admission.

**Main outcome measures:** Standardized hospitalization incidence difference and relative risk of hospitalization for a series of targeted vascular diseases from January 1 to October 31 for 2021 versus 2019. Demographic data from 2019 were used for the standardization of patient counts by 10-year age strata for each morbidity and year.

**Results:** While the relative risk of hospitalization in 2021 versus 2019 decreased for almost all diseases, an increase in relative risk was observed for myocarditis (28.0%) and pulmonary embolisms (10.0%).

In 2020, the relative risk of hospitalization versus 2019 also decreased for almost all diseases but remained stable for myocarditis and increased by 4.0% for pulmonary embolisms.

In 2021, the difference in myocarditis coincided with the vaccination campaign in young individuals. The increase in pulmonary embolism occurred predominantly in older women, with a weak but still noticeable coincidence with the vaccination campaign.

**Conclusions:** The deficit in care for patients with acute atherothrombotic manifestations in 2021 and 2020 shows a failure by the French healthcare system to rectify the deficiencies of 2020. The risk excess for pulmonary embolism cannot be entirely explained by covid-19 or by vaccine-induced immune thrombotic thrombocytopenia. This warrants investigating the risk/efficacy ratio of a temporary thromboprophylaxis in individuals at risk before vaccine.

## Introduction

Cardiovascular illnesses represent a major health burden in industrialized countries and are responsible for a large proportion of hospital bed occupancies. In 2019 in France, medical care for cardiovascular diseases (CVD) accounted for 8.6% of hospital stays, only second to digestive diseases.^1^ Variations in CVD incidence thus weigh heavily on public health management, impacting the provisional organization of healthcare services.

The three first official covid-19 cases in France were identified on January 24, 2020. The covid-19 pandemic caused a global perturbation in hospital activity with the saturation of regular and intensive care bed capacities and the postponement or cancellation of many scheduled hospitalizations (or the avoidance of unscheduled ones). In addition to social distancing, masking, curfews and travel limitations, the French government ordered three lockdowns: from March 17th to May 11th, 2020, then from October 30th to December 15th, 2020, and finally from April 3rd to May 3rd, 2021. Finally, the covid-19 vaccination campaign was launched on December 27^th^, 2020.^2^

As regards the impact of the pandemic on CVD, the global spread of covid-19 raises the risk of numerous non-pulmonary consequences, the most severe being cardiovascular problems,^3,4^ as severe covid-19 is highly thrombogenic.^5,6^ Concerning vaccination, rare side effects include myocarditis^7^ after the mRNA-BNT162b2 vaccine, and thrombotic events from vaccine immune-induced thrombotic thrombocytopenia, after vector-based vaccines.^8,9^

Our goal was thus to perform a retrospective analysis of a series of targeted cardiovascular pathologies across France, comparing the incidence of inpatient stays before versus during the covid-19 pandemic at an exhaustive, nation-wide level, giving special attention to variations observed during the covid-19 vaccination campaign.

## Methods

### Setting and data sources

A nationwide, retrospective, exhaustive, population-based cohort study was conducted using the French National Uniform Hospital Discharge Data Set Database (PMSI)^10^ for years 2019 to 2021. The latter includes all inpatient hospital stays in private and public hospitals in a standardized, anonymized data set. According to French law, no informed consent was required to use these data (due to their anonymous nature). The available data include age, sex, dates of admission and discharge, length of stay, principal diagnosis coded according to the International statistical Classification of Diseases and related health problems, 10^th^ revision (ICD10), and the Diagnostic Related Group (DRG) assigned to the stay.

Since no official cases of COVID-19 were recorded for 2019, this year was considered as the reference year when evaluating changes in disease incidence. Furthermore, case numbers for the years 2020 and 2021 were standardized for sex and age groups using the 2019 French demographic data provided by the French National Institute of Statistics and Economic Studies (INSEE).^11^

Finally, subsets of interest were juxtaposed with vaccination data retrieved from the French Ministry of Solidarity and Health website.^12^

### Study pathologies, population, and period

We focused on a selection of cardio-vascular diseases related to thromboembolic or inflammatory processes, identified by their ICD10 codes. These pathologies fall into five broad groups: (group 1) “Heart – Ischemic” with myocardial infarction and angina pectoris; (group 2) “Heart – Inflammation” for myocarditis and pericarditis; (group 3) “Cerebrovascular” for transient ischemic attack (TIA), stroke; (group 4) “Arterial embolism and thrombosis”; and (group 5) “Venous embolism and thrombosis” with deep vein thrombosis, pulmonary embolism and other venous thromboses (see eTable 1 for the related ICD10 codes).

In order to focus on incident cases, we first selected stays with one of the targeted pathologies coded as the principal diagnosis. For each pathology, we then retained only individuals hospitalized during the 2019 to 2021 period for the first time in two years. We considered these patients as incident for the pathology (year+2 incident patients).

Records with invalid identifiers or error DRG were excluded. Patients from outside Metropolitan France were also excluded, since oversea territories have specific populations and health statuses often highly influenced by singular socio-economic environments and health resource allocation (e.g. the migrant crisis on the island of Mayotte).

Furthermore, the PMSI database contains only discharged hospitalizations, resulting in a lack of stays at the end of the available period due to unfinished hospitalizations. In order to limit this bias, we computed the 95th to 99th percentiles of length of stay for the target pathologies. The percentiles of length of stay were computed for years 2019, 2020 and 2021 to control any length of stay variation due to the pandemic. Based on these data, we determined that the first ten months of each year cover at least 99% of the stays ending in December (see eTable 2).

### Study outcomes

The primary outcome was the difference in standardized incidences between 2021 and 2019. Secondary outcomes include the differences between years 2020 and 2019 and between years 2021 and 2020. In practice, we produced a hierarchical data exploration system comprising multiple summary figures per pathology (https://osf.io/2b7hk/).^13^ Starting from a main table containing the global information about each pathology, it is possible to navigate and explore a given pathology by drilling down to sex and age group.

The underlying data concerning a) caseload by sex and age group per year and b) the difference in caseloads per month (all age groups combined), are also provided (https://osf.io/2b7hk/).^13^ In order to comply with French regulations concerning privacy and medical secrecy, absolute patient numbers equal to or under 10 are replaced by ‘NA’.

### Statistical Analysis

#### General analysis

The number of patients per each 10-month period for 2020 and 2021 was standardized by sex and 10-year age strata (from 0 to 9 years to 80 years and over) based on the 2019 national population data. We calculated the expected number of patients with the associated 95% confidence interval (95% CI) had the population characteristics for sex and age strata been identical to the reference year. Bar charts were generated for each morbidity, representing case numbers and standardized case numbers by sex and age strata for 2019, 2020 and 2021.

We also computed the difference between each dyad of years, with the corresponding relative risk (RR). This analysis was also performed for each morbidity and each sex, and then sex and age strata. Statistical analyses were performed using R software (command “riskratio.wald” from package “etpitools”), with a 2-tailed significance threshold of α = 0.05.

#### Supplementary analyses

Considering the general aspect of changes in incidence between 2020 and 2019, we further focused on the impact of the first lockdown in France. Incidence differences between 2020 and 2019 for the period ranging from March 1^st^ to May 10^th^ were computed, differentiating positive and negative changes. It was thus possible to assess their respective contributions to the overall change.

Finally, for two pathologies, we juxtaposed incidence data and vaccination data (numbers of first doses injected per month in 2021), with the limitation that the vaccination data are only available by age group, not sex. To explore potentially vaccine-associated pathologies, we performed a) an additional extraction separating stays with and without a diagnostic code for symptomatic Covid (see eTable 3) as a way to eliminate cases potentially caused by Covid-19, and b) we computed the difference in the number of stays with or without Covid between 2020 and 2021. Since there was a sharp decline in stays during March 2020, we replaced the March 2020 values by the 2019 March values, applying the March 2020 Covid/Non Covid ratio.

## Results

### Main results

Overall, 1,006,819 patients were selected over the ten target pathologies. After standardization and selection of the period studied (ten first months) for each year, the population size is 919,514 (Table 1). Generally, hospitalizations for all pathologies sharply declined in the first half of March 2020 through April, preceding the first lockdown (Figure 1). Considering changes in incidence during this period (see eTable 4), five different cases can be broadly distinguished: i) more than 35% decline for arterial embolism/thrombosis and angina pectoris; ii) approximately 30% decline for other and deep venous thromboses; iii) transient ischemic attack, myocarditis, and myocardial infarction with a decline of 20 to 23%; iv) pericarditis and stroke with 13 and 14% decrease, and v) pulmonary embolism, with a mere 4% decrease being the result of a sharp drop (−19%) during the first half of March 2020 followed by a strong surge (+14%).

**Table 1.** Numbers of year+2 incident patients for the first 10 months of 2019, 2020 and 2021, standardized to the corresponding 2019 population data, with 95%CI for years 2020 and 2021.

| Morbidity | N2019 | N2020 (95%CI) | N2021 (95%CI) |
| --- | --- | --- | --- |
| Angina pectoris | 50,297 | 43,248 (42,842 to 43,658) | 42,189 (41,788 to 42,594) |
| Myocardial infarction | 70,937 | 67,789 (67,280 to 68,301) | 69,530 (69,014 to 70,049) |
| Pericarditis | 6,632 | 5,944 (5,795 to 6,098) | 5,908 (5,759 to 6,061) |
| Myocarditis | 2,098 | 2,049 (1,961 to 2,140) | 2,680 (2,580 to 2,784) |
| Transient ischemic attack | 28,977 | 26,895 (26,575 to 27,219) | 26,472 (26,155 to 26,793) |
| Stroke | 80,620 | 76,849 (76,307 to 77,394) | 77,242 (76,699 to 77,789) |
| Arterial embolism and thrombosis | 33,292 | 29,108 (28,776 to 29,445) | 30,900 (30,557 to 31,247) |
| Pulmonary embolism | 32,184 | 33,571 (33,213 to 33,932) | 35,517 (35,149 to 35,888) |
| Deep vein thrombosis | 7,980 | 6,746 (6,587 to 6,910) | 6,272 (6,118 to 6,430) |
| Other venous thromboses | 6,308 | 5,581 (5,436 to 5,730) | 5,699 (5,552 to 5,849) |

**Figure 1.**
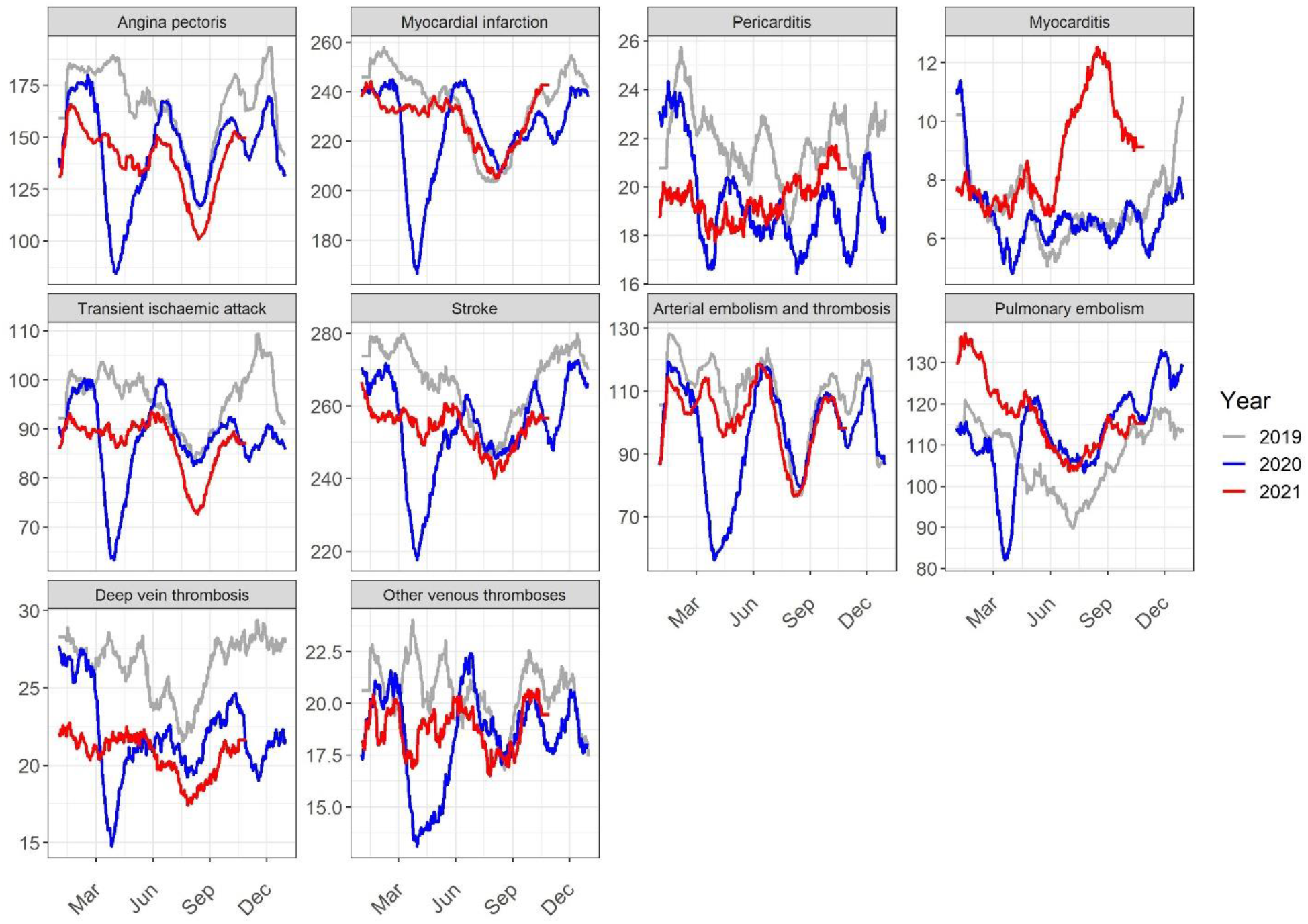
General trend (28-day sliding average) in caseloads per day for each pathology.

### Patterns in annual change

Based on a) the ratio of cases between each consecutive year and the 2021/2019 global evolution, b) the trend in cases over the years, and c) the relative risk between each consecutive year and the 2021/2019 relative risk (Table 2), we identify 4 groups of pathologies with similar temporal patterns.

**Table 2.**
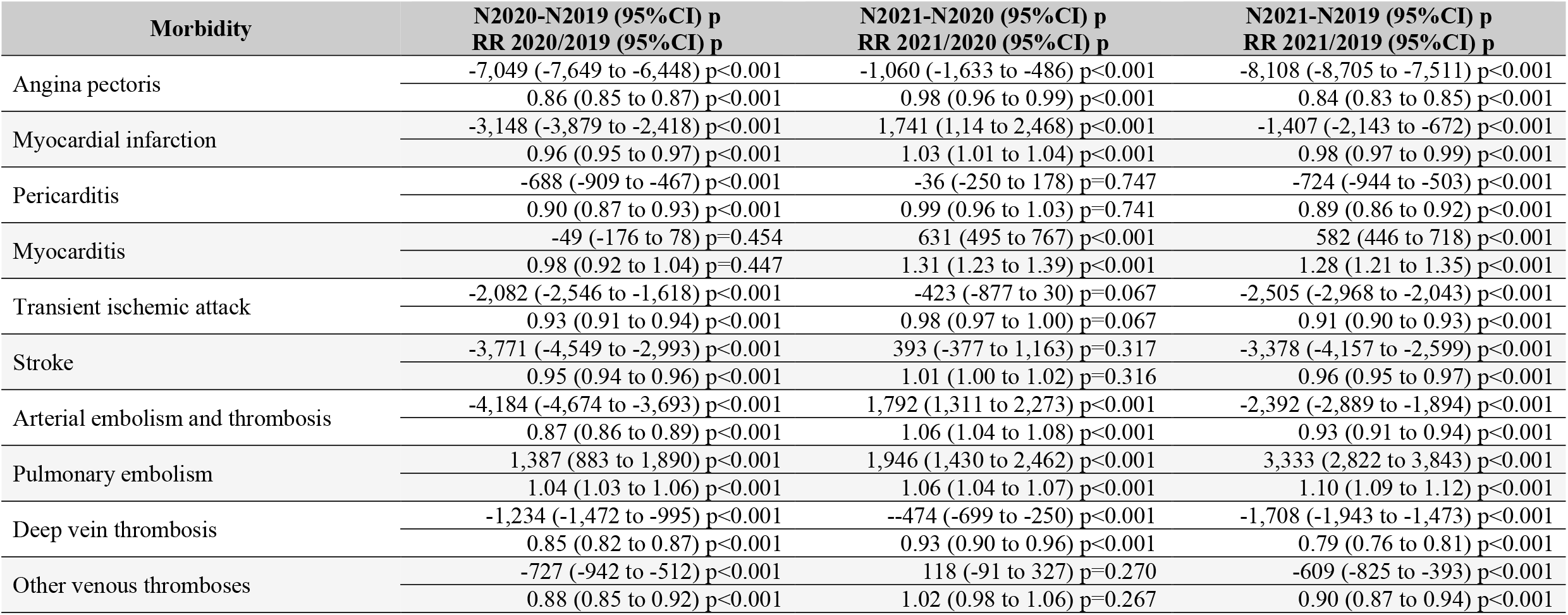
Differences in year+2 incidence and relative risk (RR X/Y: for the year X, the year Y being the reference) for the first 10 months between year dyads, with their 95%CI and statistical significance (p).

#### Group A: “Sustained decrease”

The incidence and relative risk of hospitalization significantly decrease each year and overall. This group contains deep venous thromboses (21.4% overall decrease) and angina pectoris (16.1% overall decrease).

#### Group B: “Attenuated decrease”

After a significant decrease in 2020, there is no significant change between 2020 and 2021, although the global 21/19 balance still shows a significant decrease. This is the case for pericarditis (11.6% overall decrease), other venous thromboses (9.7% decrease), transient ischemic attacks (8.6% decrease), and strokes (4.2% decrease).

#### Group C: “Partial rebound”

Significant decrease in 2020 followed by a significant increase in 2021, with an overall significant decrease. This pattern occurs for arterial embolisms and thromboses (12.6% decrease in 2020 before increasing by 6.2% in 2021, and global balance of - 7.2%) and myocardial infarctions (4.4% drop in 2020, 2.6% incidence rebound in 2021, and global balance of -2.0%).

#### Group D: “Increase”

In this group, the overall incidence and relative risk of hospitalization globally increased, independent of 20/19 changes. This group contains myocarditis and pulmonary embolism. Myocarditis incidence non-significantly decreases in 2020 and then significantly increases in 2021 by 30.8% for an overall progression of 27.7%. In contrast, pulmonary embolisms steadily increase, with a 4.3% 20/19 followed by a 5.8% increase in hospitalization incidence for an overall progression of 10.4%.

### Focus on myocarditis and pulmonary embolism

Considering the characteristics of group D, we proceeded with further analyses using the hierarchical data exploration system and the shared study dataset (https://osf.io/2b7hk/).^13^ The global case differential for myocarditis rose progressively from May to August and recedes in September and October, notably with a significant difference between 2021 and 2020 in caseload and relative risk of hospitalization from July to September: 21/20 RR of 1.63 (1.37 to 1.95), 1.86 (1.57 to 2.20), and 1.68 (1.40 to 2.02). Compared to 2019, myocarditis occurs more frequently in 2021 among men in the 10-19 and 20-29 year age groups (Figure 2).

**Figure 2.**
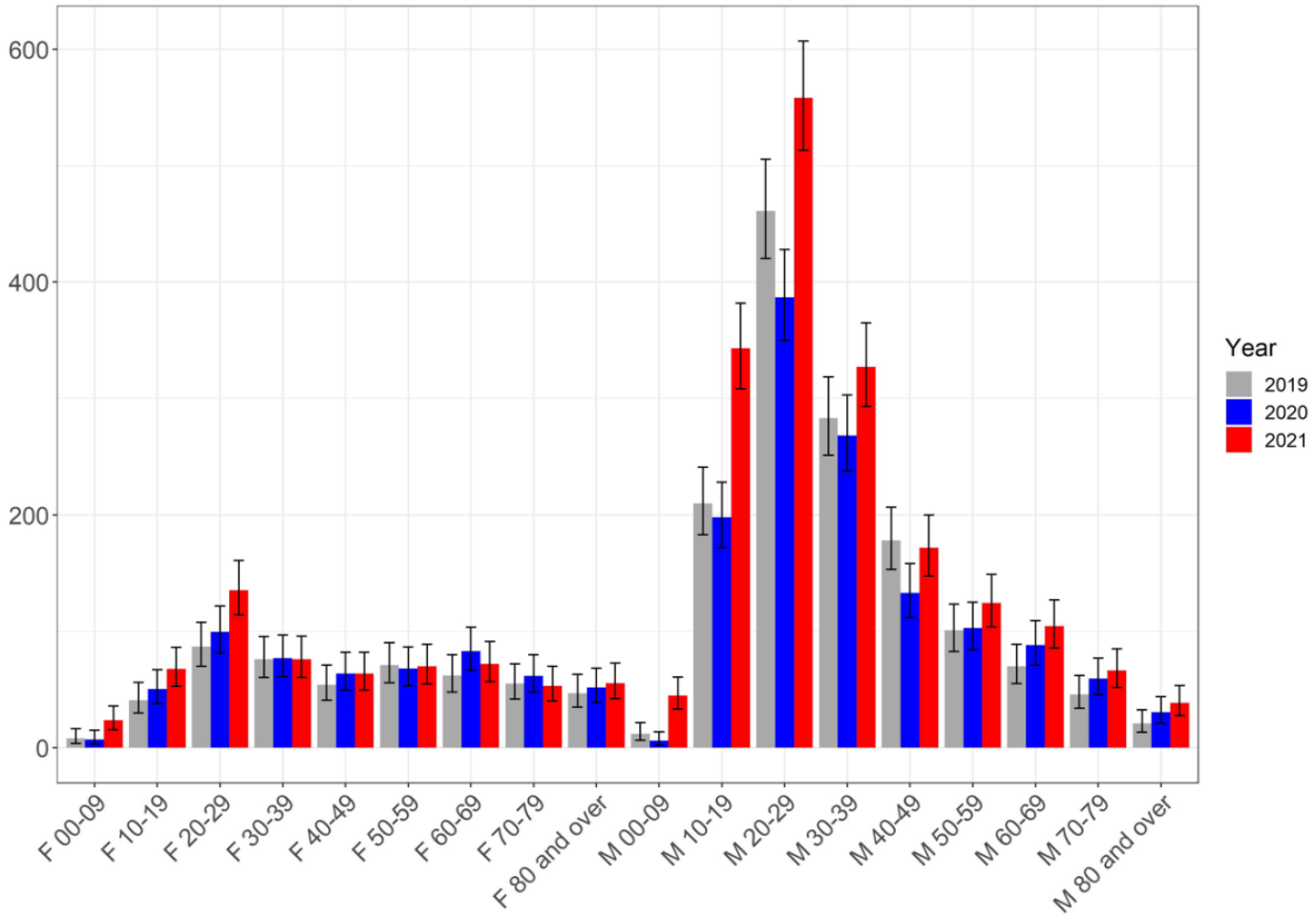
Myocarditis caseload per sex, age, and year.

An entirely different situation was observed for the pulmonary embolism group: the caseload excess is more smoothly distributed over sex-age groups with significant differences concentrated on the oldest patients, especially among women 70 and over (Figure 3).

**Figure 3.**
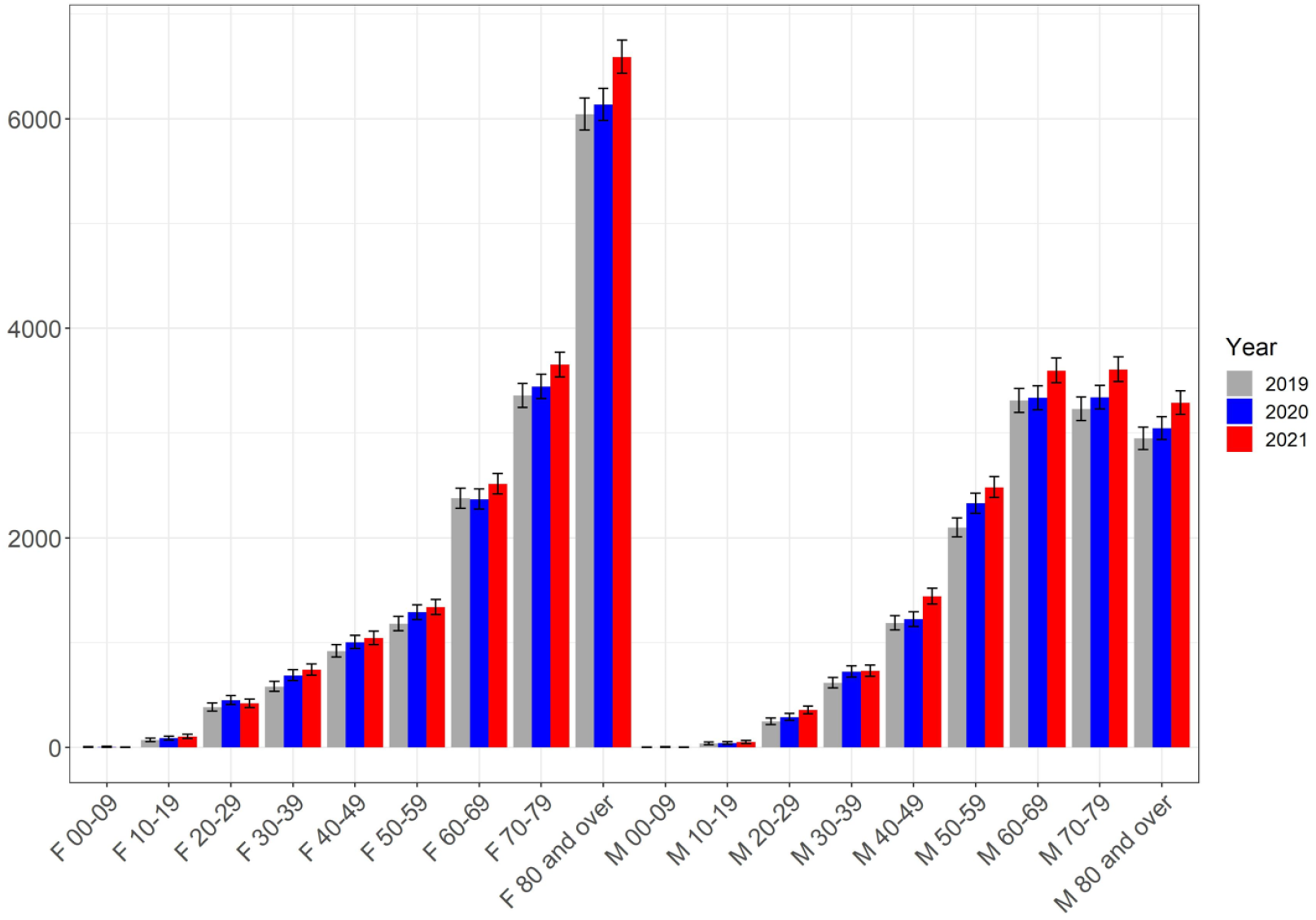
Pulmonary embolism caseload per sex, age, and year.

Results were further juxtaposed with vaccination data. In the case of myocarditis, for men aged 10 to 29, we notice a significant increase in non Covid-19 stays between 2020 and 2021 (NCS20-21) in March and then from June to October (for a total of 456 cases), following the curve of the first dose of vaccine injections for this age group (Figure 4). For pulmonary embolism among women aged 70 and over, we find a similar trend between NCS20-21 and the number of first dose injections (Figure 5), notably with a significant increase of 252 cases in April.

**Figure 4.**
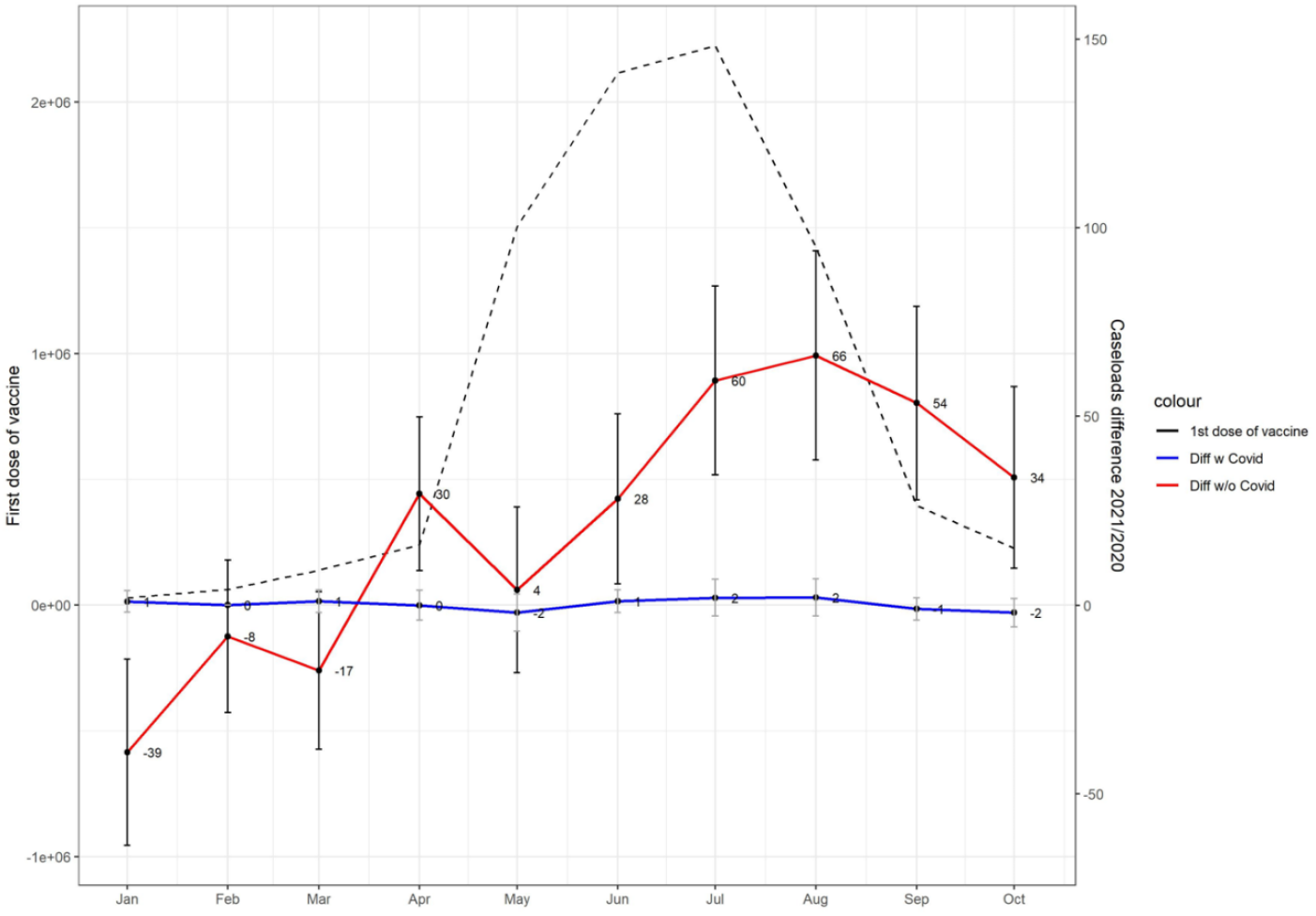
Myocarditis cases and vaccination. Difference in cases with or without Covid between 2020 and 2021, men aged 10 to 29, put in perspective with the first-dose-of-vaccination curve

**Figure 5.**
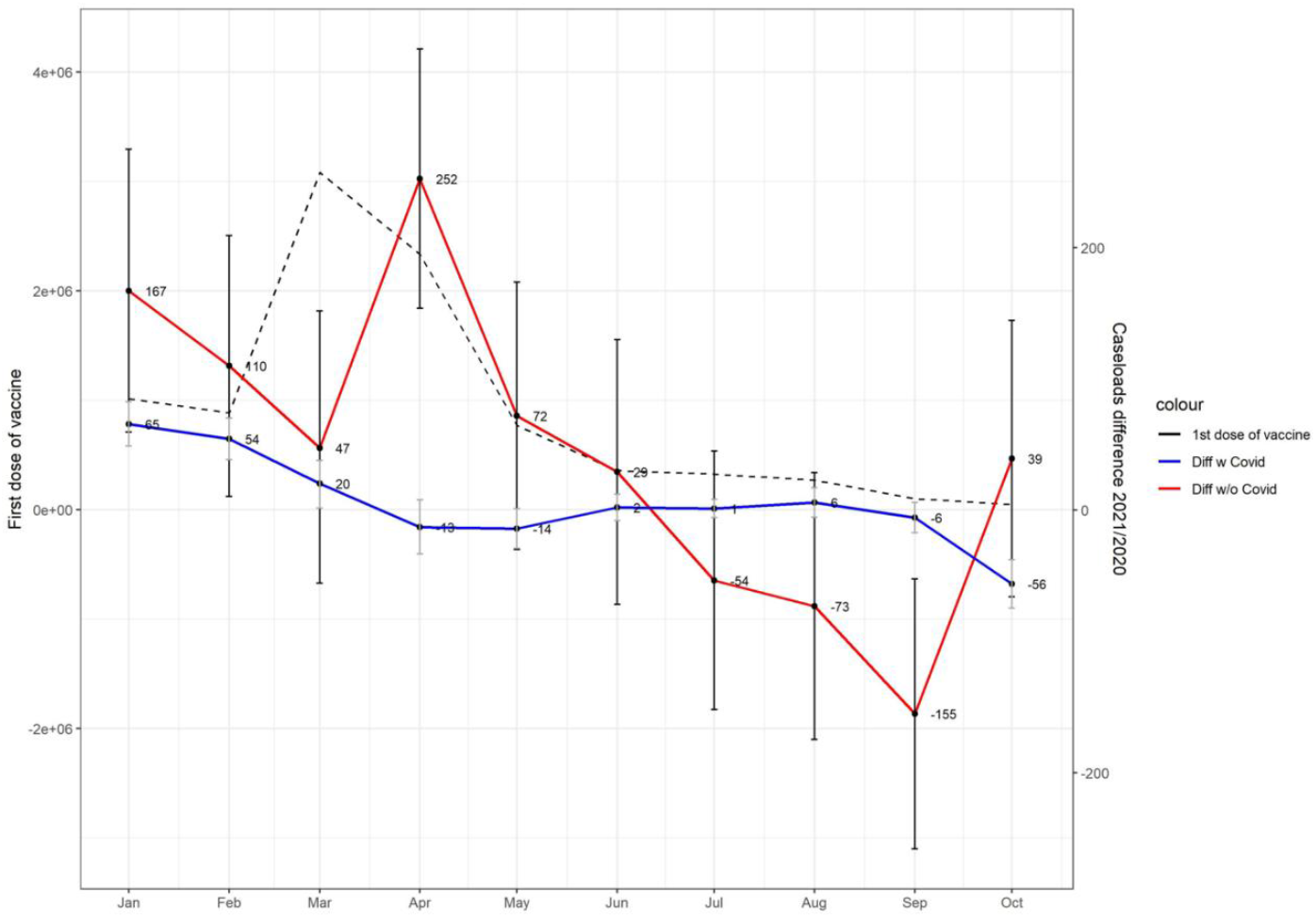
Pulmonary embolism cases and vaccination. Difference in cases with or without Covid between 2020 and 2021, women aged 70 and over, put in perspective with the first-dose-of vaccination curve.

## Discussion

### General findings

All target cardiovascular pathologies decreased in incidence for the first ten months of 2021 compared to 2019, with the exceptio of myocarditis and pulmonary embolism. For these two pathologies, incident hospitalizations significantly increased in both 2020 and 2021.

### Lessons not learned

Initial case reports and cohort studies linked covid-19 with an increased risk of acute myocardial infarction and stroke.^14–16^ This has been confirmed in a nationwide register-based study of all patients with covid-19,^17^ however reports in Denmark as well as the United States showed a strong decline in hospitalization for myocardial infarction and cardiac catheterization during the early phase of the covid-19 pandemic mitigation measures.^18,19^ In particular, the covid-19 pandemic adversely affects timely access to cardiac catheterization services.^20^ Furthermore, admissions for acute myocardial infarction in Italy were significantly reduced, with a parallel increase in fatality and complication rates.^21^ This presents an alarming picture whereby a substantial proportion of patients did not access hospital services, with a marked decrease in hospital admissions before and during the first lockdown in France.^22^ Our data support this hypothesis, demonstrating approximate declines of 36% and 20% in hospitalization incidence for angina pectoris and myocardial infarction, respectively, in March/April 2020 compared to 2019. Thus, the comparative increase in hospitalizations for acute coronary atherothrombosis in 2021 is probably a partial re-normalization of the necessary hospital care, still insufficient when comparing the rates for 2021 to those for 2019.

Similarly, stroke neurologists from Italy, France, and Germany observed limited or delayed access to acute stroke diagnostics and time-dependent therapies.^23^ The general observation is therefore that of a global restriction of hospital care channels for acute atherothrombotic, coronary and cerebrovascular pathologies induced by the covid-19 pandemic. This suggests that the French healthcare system struggled to provide care in 2021, despite the lessons learned in 2020.

### Myocarditis

Like other viral illnesses,^24^ acute myocarditis is associated with covid-19.^25–27^ It is most often associated with symptomatic covid-19, which is by itself the reason for hospitalization. In the setting of our study, it is therefore unsurprising that we did not observe excess myocarditis cases in 2020; on the contrary, myocarditis incidence declines by 20% circa the first lockdown. However, we cannot exclude an increase in the number of cases with a predominant myocarditis masked by the difficulty to access hospital cardiology channels. An excess risk of myocarditis has been previously described in patients receiving at least one dose of the mRNA vaccines against SARS-CoV-2, referring to both the Spikevax® mRNA-1273 vaccine and the Comirnaty® BNT162b2 vaccine.^28–30^ This was also observed in France in the VigiBase national database of adverse drug reaction reports.^31^ Adverse vaccine reactions are thus likely to explain at least a part of the myocarditis case excess observed in 2021 compared to 2020 in the present study. Taking into account available data for the vaccination campaign for people aged 10 to 29, we note that the increase in myocarditis cases in 2021 relative to 2020 for men in this age group coincides with the curve for the first dose of vaccine.

### Pulmonary embolism

A French nationwide study reported an increased number of patients hospitalized with pulmonary embolism in 2020, mainly linked to the covid-19 waves, in both patients with and without covid-19.^32^ This increase was also observed in the United Kingdom,^33^ but not in Denmark.^34^ In the present study, the difference between the 2021 and 2020 periods mainly occurred in the first five months, and was concentrated in non-covid-19 patients. This effect persisted even after conservatively compensating for a probable under-representation of cases in 2020. Furthermore, the first months in 2021 coincided with the beginning of the vaccination campaign in France, and we observe a co-occurrence between the increase in pulmonary embolism cases in 2021 versus 2020 for women aged 70 and over, and the curve of the first dose of vaccine for people aged 70 and over. We therefore suggest an increased risk of pulmonary embolism associated with the vaccination campaign.

Since the rollout of the covid-19 vaccine, there have been reports of rare venous thrombosis such as cerebral venous sinus and portal/splanchnic/hepatic veins thrombosis, mostly linked to the vaccine-induced immune thrombotic thrombocytopenia^8,9,35,36^ following the ChAdOx1 nCoV-19 vaccine. One case of isolated pulmonary embolism associated with thrombocytopenia was described among the 11 cases of the initial Geinacher’s report^9^ and another case later.^36^ Very few cases of isolated pulmonary embolism^37^ or deep vein thrombosis^38^ exist in the literature after covid-19 vaccination in the absence of a blood cell count abnormality. A recently available meta-analysis of the risk of vaccine-induced thrombotic thrombocytopenia (VITT) following ChAdOx1 nCoV-19 vaccine found a pooled incidence of 7.3 [95% CI 4.3 to 12.3] per 1,000,000 persons receiving a first dose of ChAdOx1 nCoV-19 vaccine compared with the reported incidence in France of 6.3 [95% CI 2.9 to 1.19].^41^ Thus only a fraction of the excess cases of pulmonary embolism found during the first ten months of 2021 can be attributed to VITT alone.

While the BNT162b2 mRNA vaccine was the first authorized vaccine and most widely used in older persons in France, a recent French population-based study on the risk of severe cardiovascular events among people aged 75 years or older found no increase in the incidence of pulmonary embolism in the 14 days following each BNT162b2 mRNA vaccine dose.^42^ However, as some pharmacokinetic studies consider post-vaccine peak immunity to occur after 14 days,^43^ there might be a selection bias in the time frames considered.

### Strength and limitations

The strength of our study is its nationwide scale, based on the systematic coding of diseases in hospitalized patients, strongly supporting generalizability to the French population. The data are subject to quality controls led by the French Social Security administration, potentially resulting in fines, thus ensuring a certain level of accuracy.

However, there are two main limitations. First is possible misclassification, potentially generating strong biases. Second, we do not have any information on outpatients and patients treated at home, and for example, we cannot assess a decrease in hospitalization rates for diseases linked with disease-related deaths at home. We also do not have any access to the type of vaccine administered and can only take note of the temporal concordance between case excess and vaccination.

### Conclusion

The covid-19 pandemic in 2021 was associated with a deficit in care for patients with acute atherothrombotic manifestations. These issues were a repeat of those seen in 2020, suggesting a failure of the French healthcare system to adapt to pandemic conditions. In parallel with the vaccination campaign, an excess risk of myocarditis was confirmed in 2021. The 2021 period was also associated with an excess risk of pulmonary embolism, which cannot be entirely explained by vaccine-induced immune thrombotic thrombocytopenia. This result warrants further investigation, in order to determine the risk/efficacy ratio of a temporary thromboprophylaxis in individuals at significant risk of venous thromboembolism.

## Supporting information

RECORD checklist

## Data Availability

All data produced are available online at https://osf.io/2B7HK/

## Acknowledgements

We thank Sarah Kabani, medical writer (BESPIM), for editing the manuscript, which was funded by the CHU Nîmes in accordance with GPP3 guidelines.

## Author’s contributions

TB is guarantor for this manuscript and had full access to all of the data in the study and takes responsibility for the integrity of the data and the accuracy of the data analysis. Concept and design of the study: TB, LG, J-CG. Acquisition, analysis, or interpretation of data: TB, LG, AP-M, CMS, J-CG. Drafting of the manuscript: J-CG, TB. Critical revision of the manuscript for important intellectual content: TB, LG, AP-M, CMS, J-CG. Statistical analysis: TB, LG. Administrative, technical, or material support: TB, LG. Supervision: J-CG, TB.

## Funding

This research project was supported by internal funding from the Nimes University Hospital (the study sponsor).

## Data sharing

PMSI data are available for researchers who meet the criteria for access to these confidential data (this access is submitted to the approval of the National Committee for data protection) from the national agency for the management of hospitalisation (ATIH – Agence Technique de l’Information sur l’Hospitalisation). We are not allowed to share the raw data and provide aggregated data, replacing values equal to or under 10 by ‘NA’. We also provide a hierarchical data exploration system comprising multiple summary figures per pathology https://osf.io/2b7hk/.

## Ethics approval and consent to participate

The study was approved by the Institutional Review Board of the University Hospital of Nîmes, reference number 21.12.28. Retrospective studies on the National Uniform Hospital Discharge Data Set Database do no need legal authorization, and written consent is not needed since data are anonymous. Written consent was not needed for this study as it is a retrospective study and the national data used are anonymous.

## Competing interests

Carey Suehs reports grants and personal fees from Astra Zeneca, grants from GSK, unrelated to the current work. The remaining authors report no competing interests.

## Notes

### Funding Statement

This study was supported by internal funding from the Nimes University Hospital (the study sponsor).

### Author Declarations

The Institutional Review Board of the University Hospital of Nimes gave ethical approval for this work.

### Summary of Updates

Figures from the supplement are integrated directly in the main manuscript. A bug in the generation of one figure was corrected. All displays are now inline with the text. Minor typos were corrected.

