## Supplementary tables and figures for "Change in incidence of cardiovascular diseases during the covid-19 pandemic and vaccination campaign: data from the nationwide French hospital discharge database"

### Supplemental material

eTable 1 – ICD10 codes used to identify the targeted cardiovascular pathologies (when used as the principal diagnosis)

eTable 2 – Percentile of length of stay for the targeted pathologies

eTable 3 – ICD10 codes used to identify stays with Covid (used as principal or secondary diagnosis)

eTable 4 – Comparison of case numbers between 2019 and 2020 for the period running from March 1st to May 10th, differentiating positive and negative changes

Hierarchical Data Exploration System

Data model for data provided

**eTable1**

| Group | Pathology | Principal Diagnosis |
|---|---|---|
| Heart – Ischemic | ANG – Angina pectoris | I20 Angina pectoris |
| | INF – Myocardial infarction | I21 Acute myocardial infarction |
| Heart – Inflammation | MYOC – Myocarditis | I40 Acute myocarditis |
| | PERIC – Pericarditis | I30 Acute pericarditis |
| Cerebrovascular | TIA – Transient ischaemic attack | G45 Transient cerebral ischaemic attacks and related syndromes |
| | STROKE – Stroke | I61 Intracerebral haemorrhage<br>I63 Cerebral infarction<br>I64 Stroke, not specified as haemorrhage or infarction |
| Arterial embolism and thrombosis | THRB_EMB_ART – Arterial embolism and thrombosis | I74 Arterial embolism and thrombosis |
| Venous embolism and thrombosis | DVT – Deep vein thrombosis | I801 Phlebitis and thrombophlebitis of femoral vein<br>I802 Phlebitis and thrombophlebitis of other deep vessels of lower extremities<br>I822 Embolism and thrombosis of vena cava |
| | EMB – Pulmonary embolism | I26 Pulmonary embolism |
| | OTH_VT – Other venous thromboses | I803 Phlebitis and thrombophlebitis of lower extremities, unspecified<br>I808 Phlebitis and thrombophlebitis of other sites<br>I809 Phlebitis and thrombophlebitis of unspecified site<br>I828 Embolism and thrombosis of other specified veins<br>I829 Embolism and thrombosis of unspecified vein |

**eTable 1 – ICD10 codes used to identify the targeted cardiovascular pathologies (when used as the principal diagnosis)**

**eTable2**

| TypePath | Year | # stays | Q95 | Q96 | Q97 | Q98 | Q99 | Q100 |
|---|---|---|---|---|---|---|---|---|
| ANG – Angina pectoris | 2019 | 60,387 | 8 | 9 | 11 | 13 | 17 | 316 |
| | 2020 | 53,163 | 8 | 9 | 10 | 12 | 16 | 117 |
| | 2021 | 51,689 | 8 | 8 | 10 | 12 | 15 | 118 |
| INF – Myocardial infarction | 2019 | 86,059 | 16 | 18 | 20 | 23 | 29 | 391 |
| | 2020 | 83,204 | 15 | 17 | 19 | 22 | 28 | 182 |
| | 2021 | 85,544 | 15 | 16 | 18 | 21 | 27 | 217 |
| PERIC – Pericarditis | 2019 | 7,956 | 14 | 15 | 18 | 21 | 26 | 98 |
| | 2020 | 7,121 | 14 | 16 | 18 | 21 | 27 | 113 |
| | 2021 | 7,237 | 15 | 17 | 19 | 22 | 28 | 129 |
| MYOC – Myocarditis | 2019 | 2,579 | 12 | 14 | 16 | 18 | 26 | 123 |
| | 2020 | 2,481 | 14 | 15 | 19 | 23 | 31 | 114 |
| | 2021 | 3,325 | 12 | 14 | 15 | 19 | 26 | 215 |
| TIA – Transient ischaemic attack | 2019 | 35,176 | 11 | 13 | 14 | 16 | 20 | 119 |
| | 2020 | 32,749 | 11 | 12 | 14 | 15 | 20 | 109 |
| | 2021 | 32,281 | 11 | 12 | 13 | 15 | 20 | 124 |
| STROKE – Stroke | 2019 | 97,574 | 29 | 33 | 37 | 43 | 55 | 488 |
| | 2020 | 94,786 | 29 | 32 | 36 | 43 | 56 | 747 |
| | 2021 | 96,022 | 30 | 33 | 37 | 44 | 55 | 386 |
| THRB_EMB_ART – Arterial embolism and thrombosis | 2019 | 39,573 | 20 | 22 | 26 | 31 | 41 | 458 |
| | 2020 | 35,736 | 19 | 22 | 25 | 30 | 42 | 445 |
| | 2021 | 37,666 | 19 | 22 | 25 | 31 | 42 | 216 |
| EMB – Pulmonary embolism | 2019 | 39,226 | 20 | 21 | 23 | 27 | 33 | 197 |
| | 2020 | 41,715 | 19 | 21 | 23 | 27 | 33 | 226 |
| | 2021 | 44,200 | 20 | 21 | 24 | 27 | 34 | 263 |
| DVT – Deep vein thrombosis | 2019 | 9,682 | 17 | 19 | 21 | 24 | 30 | 182 |
| | 2020 | 8,155 | 18 | 20 | 22 | 25 | 33 | 121 |
| | 2021 | 7,737 | 17 | 19 | 21 | 24 | 31 | 114 |
| OTH_VT – Other venous thromboses | 2019 | 7,529 | 14 | 15 | 18 | 20 | 25 | 90 |
| | 2020 | 6,798 | 13 | 14 | 16 | 20 | 25 | 141 |
| | 2021 | 6,991 | 13 | 15 | 17 | 20 | 27 | 127 |

**eTable 2 – Percentile of length of stay for the targeted pathologies**

**eTable3**

| Code | Label |
|---|---|
| **U0710** | COVID-19, respiratory symptoms with identification |
| **U0711** | COVID-19, respiratory symptoms without testing |
| **U0714** | COVID-19, other symptoms with identification |
| **U0715** | COVID-19, other symptoms without testing |

### eTable 3 – ICD10 codes used to identify stays with Covid (used as principal or secondary diagnosis)

**eTable4**

| Morbidity | N2020-N2019 (95%CI) p | (N2020-N2019)/N2019 (95%CI) p |
|---|---|---|
| Angina pectoris | -4,598 (-4,881 to -4,316) p<0.001 | -36% (-39% to -34%) p<0.001 |
| Myocardial infarction | -3,177 (-3,524 to -2,831) p<0.001 | -19% (-21% to -17%) p<0.001 |
| Pericarditis | -220 (-326 to -114) p<0.001 | -14% (-21% to -7%) p<0.001 |
| Myocarditis – Neg | -107 (-165 to -48) p<0.001 | -22% (-34% to -10%) p<0.001 |
| Myocarditis – Pos | 2 (-18 to 22) p=0.941 | 4% (-40% to 48%) p=0.941 |
| Myocarditis – Tot | -105 (-167 to -44) p<0.001 | -20% (-31% to -8%) p<0.001 |
| Transient ischemic attack | -1,630 (-1,851 to -1,409) p<0.001 | -23% (-26% to -20%) p<0.001 |
| Stroke | -2,569 (-2,941 to -2,197) p<0.001 | -13% (-15% to -11%) p<0.001 |
| Arterial embolism and thrombosis | -2,944 (-3,169 to -2,719) p<0.001 | -37% (-40% to -34%) p<0.001 |
| Pulmonary embolism – Neg | -786 (-959 to -613) p<0.001 | -19% (-23% to -14%) p<0.001 |
| Pulmonary embolism – Pos | 484 (317 to 652) p<0.001 | 14% (9% to 19%) p<0.001 |
| Pulmonary embolism – Tot | -301 (-542 to -61) p=0.014 | -4% (-7% to -1%) p=0.014 |
| Deep vein thrombosis | -564 (-677 to -450) p<0.001 | -29% (-35% to -23%) p<0.001 |
| Other venous thromboses | -455 (-556 to -354) p<0.001 | -30% (-36% to -23%) p<0.001 |

### eTable 4 – Comparison of case numbers between 2019 and 2020 for the period running from March 1st to May 10th, differentiating positive and negative changes

**Hierarchical Data Exploration System**
The hierarchical data exploration system (HDES) is a set of figure files navigable through html files. It is available as a compressed file (hierarchical_data_exploration_system.7z) for download at https://osf.io/2b7hk/ .

**Data model for data provided**
Two files (sex_age_year.csv and diff_month.csv) are provided, available for download at https://osf.io/2b7hk/ (data.7z).

**1. sex_age_year.csv -** Number of patients per sex and age group per year.

| Field name | Description |
|---|---|
| Patho | Pathology |
| Sex | Sex |
| Age | Age group |
| Nb2019 | Number of patients for Sex and Age Group in 2019 |
| Nb2020 | Standardized number of patients for Sex and Age Group in 2020 |
| Nb2020_min | Standardized number of patients for Sex and Age Group in 2020, lower bound |
| Nb2020_max | Standardized number of patients for Sex and Age Group in 2020, upper bound |
| Nb2021 | Standardized number of patients for Sex and Age Group in 2021 |
| Nb2021_min | Standardized number of patients for Sex and Age Group in 2021, lower |
| Nb2021_max | Standardized number of patients for Sex and Age Group in 2021, upper bound |

Then a series of fields named according the following X_Y pattern, X and Y being the years compared. For instance:

| | |
|---|---|
| Diff_X_Y | Number of patients year Y - Number of patients year X |
| Diff_X_Y_min | Number of patients year Y - Number of patients year X, lower bound |
| Diff_X_Y_max | Number of patients year Y - Number of patients year X, upper bound |
| Diff_X_Y_p | Number of patients year Y - Number of patients year X, significance |
| Diff_X_Y_Dir | If p is significant, sign of the difference (- or +), else 0 |
| RR_X_Y | Relative Risk of hospitalization on year Y compared to year X |
| RR_X_Y_min | Relative Risk of hospitalization on year Y compared to year, lower bound |
| RR_X_Y_max | Relative Risk of hospitalization on year Y compared to year, upper bound |
| RR_X_Y_p | Relative Risk of hospitalization on year Y compared to year, significance |
| RR_X_Y_Dir | If p is significant, impact of the RR (- or +), else 0 |

Example:
Diff_19_20 – Number of patients year 2020 - Number of patients year 2019
RR_19_2020 – Relative Risk of hospitalization on year 2020 compared to year 2019

**2. diff_month.csv -** Patients per sex, age group and month: Number per year and difference and RR between each dyad of years

| Field name | Description |
|---|---|
| Patho | Pathology |
| Sex | Sex |
| Month | Month of admission |
| Nb2019 | Number of patients for Sex and Month in 2019 |
| Nb2020 | Standardized number of patients for Sex and Month in 2020 |
| Nb2020_min | Standardized number of patients for Sex and Month in 2020, lower bound |
| Nb2020_max | Standardized number of patients for Sex and Month in 2020, upper bound |
| Nb2021 | Standardized number of patients for Sex and Month in 2021 |
| Nb2021_min | Standardized number of patients for Sex and Month in 2021, lower |
| Nb2021_max | Standardized number of patients for Sex and Month in 2021, upper bound |

Then a series of fields named according to the following X_Y pattern, X and Y being the years compared. For instance:

| | |
|---|---|
| Diff_X_Y | Number of patients year Y - Number of patients year X |
| Diff_X_Y_min | Number of patients year Y - Number of patients year X, lower bound |
| Diff_X_Y_max | Number of patients year Y - Number of patients year X, upper bound |
| Diff_X_Y_p | Number of patients year Y - Number of patients year X, significance |
| Diff_X_Y_Dir | If p is significant, sign of the difference (- or +), else 0 |
| RR_X_Y | Relative Risk of hospitalization on year Y compared to year X |
| RR_X_Y_min | Relative Risk of hospitalization on year Y compared to year, lower bound |
| RR_X_Y_max | Relative Risk of hospitalization on year Y compared to year, upper bound |
| RR_X_Y_p | Relative Risk of hospitalization on year Y compared to year, significance |
| RR_X_Y_Dir | If p is significant, impact of the RR (- or +), else 0 |

Example:
Diff_19_20 – Number of patients year 2020 - Number of patients year 2019
RR_19_2020 – Relative Risk of hospitalization on year 2020 compared to year 2019
